# Role of *DOCK8* in Hyper-inflammatory Syndromes

**DOI:** 10.1101/2024.02.20.24303049

**Authors:** Mingce Zhang, Remy R. Cron, Niansheng Chu, Junior Nguyen, Scott M. Gordon, Esraa M. Eloseily, T. Prescott Atkinson, Peter Weiser, Mark R. Walter, Portia A. Kreiger, Scott W. Canna, Edward M. Behrens, Randy Q. Cron

## Abstract

**Background:** Cytokine storm syndromes (CSS), including hemophagocytic lymphohistiocytosis (HLH), are increasingly recognized as hyper-inflammatory states leading to multi-organ failure and death. Familial HLH (FHL) in infancy results from homozygous genetic defects in perforin-mediated cytolysis by CD8 T-lymphocytes and natural killer (NK) cells. Later onset CSS are frequently associated with heterozygous defects in FHL genes, but genetic etiologies for most are unknown. We identified rare *DOCK8* variants in CSS patients.

**Objective:** We explore the role of CSS patient derived *DOCK8* mutations on cytolytic activity in NK cells. We further study effects of *Dock8^-/-^* in murine models of CSS.

**Methods:** *DOCK8* cDNA from 2 unrelated CSS patients with different missense mutations were introduced into human NK-92 NK cells by foamy virus transduction. NK cell degranulation (CD107a), cytolytic activity against K562 target cells, and interferon-gamma (IFNγ) production were explored by flow cytometry (FCM). A third CSS patient *DOCK8* mRNA splice acceptor site variant was explored by exon trapping. *Dock8^-/-^* mice were assessed for features of CSS (weight loss, splenomegaly, hepatic inflammation, cytopenias, and IFNγ levels) upon challenge with lymphochoriomeningitic virus (LCMV) and excess IL-18.

**Results:** Both patient *DOCK8* missense mutations decreased cytolytic function in NK cells in a partial dominant-negative fashion *in vitro*. The patient *DOCK8* splice variant disrupted mRNA splicing *in vitro*. *Dock8*^-/-^ mice tolerated excess IL-18 but developed features of CSS upon LCMV infection.

**Conclusion:** Mutations in *DOCK8* may contribute to CSS-like hyper-inflammatory states by altering cytolytic function in a threshold model of disease.

**Key Messages:**

- Heterozygous missense mutations in *DOCK8* may contribute to decreased NK cell function via partial dominant-negative effects on perforin-mediated cytolysis.
- Heterozygous mutations in *DOCK8* may contribute to hyper-inflammatory syndromes in a threshold model of disease.
- LCMV infection of *Dock8*^-/-^ mice recapitulates features of murine FHL.

**Capsule Summary:** Heterozygous missense and splice site mutations in *DOCK8* may contribute to hyper-inflammation in patients with CSS. DOCK8 is important for optimal NK cell cytolytic function, and LCMV infection of *Dock8*^-/-^ mice resembles murine FHL.

## INTRODUCTION

Hyper-inflammatory syndromes, including hemophagocytic lymphohistiocytosis (HLH) and macrophage activation syndrome (MAS), are frequently fatal conditions resulting from a pro-inflammatory cytokine storm.(1) Familial HLH (FHL) is rare (approximately 1 in 50,000 live births) typically occurring within the first year of life, but secondary forms of HLH (sHLH) may affect up to 1 in 3,000 individuals at any age.(2) FHL results from homozygous autosomal recessive defects in proteins critical perforin-mediated cytolysis by cytotoxic CD8 T-lymphocytes and natural killer (NK) cells.(3) sHLH and MAS are associated with intracellular pathogens (e.g. Epstein-Barr virus (EBV)), hematologic malignancies (e.g. T-cell leukemia), and autoimmune (e.g. systemic lupus erythematosus (SLE)) or autoinflammatory conditions (e.g. systemic juvenile idiopathic arthritis (sJIA)).(4) The distinction between FHL and sHLH is becoming blurred as heterozygous hypomorphic or dominant-negative mutations in FHL genes are being increasingly identified as disrupting NK cell function(5) and contributing to sHLH pathogenesis using a threshold model of disease.(6) As reported, up to 20-40% of sHLH cohorts, possess heterozygous defects in known HLH genes.(7), yet a large percentage of sHLH individuals without known genetic risk factors develop a cytokine storm syndrome (CSS). During the severe acute respiratory syndrome coronavirus 2 (SARS-CoV-2) pandemic up to 20% of infected individuals developed hyper-inflammation requiring hospitalization, and a significant percentage of these hospitalized coronavirus disease 2019 (COVID-19) patients have been identified with heterozygous defects in known FHL genes, but many have not.(8)

Genetic defects in FHL genes as well as in genes associated with primary immunodeficiency (PID) and dysregulated immune activation and proliferation have been found in children with HLH.(9); in this large cohort an affected child was identified with biallelic disruption of the PID gene DOCK8 (dedicator of cytokinesis 8), not previously associated with HLH.(9)

Homozygous defects in DOCK8 are known to result in a form of autosomal-recessive Hyper-IgE syndrome.(10) DOCK8 deficiency may contribute to HLH via disruption of NK cell cytolytic function as demonstrated *in vitro*.(11, 12) DOCK8 is a GTPase important for cytoskeletal trafficking of granules, including perforin containing cytolytic granules. DOCK8 also interacts with CDC42, a product of another disrupted gene recently identified in patients with HLH(13),it plays multiple roles beyond cytotoxicity, including immune cell migration/trafficking, proliferation, and survival. We propose that heterozygous DOCK8 mutations may contribute to CSS in sHLH by partially disrupting lymphocyte cytolytic function similar to reports of complete or partial dominant-negative effects on NK cell function resulting from heterozygous mutations in other FHL associated genes identified in sHLH patients,(5, 14–17). Thus, DOCK8 should be considered as a novel HLH-associated gene.

## METHODS

### Human subjects

Children with HLH (fulfilling HLH-2004 and/or HScore criteria)(18, 19) hospitalized at Children’s of Alabama (Birmingham, AL, USA) have frequently, but not routinely, undergone commercial (GeneDx, Gaithersburg, MD, USA; and Invitae, San Francisco, CA, USA) DNA (exome) sequencing to identify FHL and associated PID genes since September 2007. Between August 2014 and September 2018, 3 of 37 children who underwent DNA sequencing for HLH were noted to have heterozygous mutations in DOCK8 (Table 1). Upon informed signed consent of subjects/families of a University of Alabama at Birmingham (UAB) Institutional Review Board (IRB) approved protocol, patient peripheral blood mononuclear cell (PBMC) derived mRNA was converted to cDNA to confirm missense mutations by Sanger sequencing, and the splice acceptor site variant was confirmed by targeted genomic DNA sequencing as previously described.(17)

**Table 1.** Demographics, diagnoses, and genetics of HLH patients with heterozygous DOCK8 mutations.

| <u>#</u> | <u>Age</u><sup>*</sup> | <u>Sex</u> | <u>Disease</u> | <u>Trigger</u> | <u>Mutation</u> | <u>Frequency</u> | <u>Pathogenic</u><sup>#</sup> |
| --- | --- | --- | --- | --- | --- | --- | --- |
| 1 | Teen | M | Asthma | Bartonella | c.782C>T, p.Ala261Val | novel | likely |
| 2 | Teen | M | JIA/ERA | ?Flare | c.4850A>G, p.Gln1617Arg | 0.1% | possibly |
| 3 | Teen | M | T-cell leukemia | ?Candida | c.54-1G>T (splice acceptor) | 0.03% | likely |

\* - 13-19 years; <sup>#</sup> - PolyPhen-2
ERA = enthesitis related arthritis; JIA = juvenile idiopathic arthritis

### DNA expression constructs

Wild-type (WT) human DOCK8 expression cDNA was cloned from healthy donor derived PBMC mRNA using reverse transcriptase (ThermoFisher, Waltham, MA, USA) as previously described.(20) Sanger sequence analysis demonstrated the cDNA sequence was equivalent to DOCK8 mRNA variant 1 (NM_203447). A recombinant Foamy virus (FV) expression system required for more efficient introduction of larger (>5kb) cDNA constructs was kindly provided by Dr. Grant Trobridge at Washington State University (Pullman, WA, USA).(21) This expression system contains the pFV-SGW plasmid expressing the exogenous gene plus enhanced green fluorescent protein (EGFP) driven by the SFFV promoter, as well as 3 helper plasmids that separately produce gag, pol, and env proteins for recombinant viral particle assembly. WT DOCK8 cDNA was inserted into pFV-SGW to generate the pFV-SGW-DOCK8-WT plasmid.(20) Based on the WT plasmid, the precise HLH patient derived mutant DOCK8 sequence equivalent cDNAs were generated using a QuikChange Lightning Site-Directed Mutagenesis Kit per the supplied instructions (Agilent Technologies, Santa Clara, CA, USA). The 2 unique patient-derived mutant cDNA were confirmed by Sanger DNA sequence analysis. Recombinant FV preparation and infection of human NK-92 NK cells was conducted as previously described.(20)

### NK cell cytolytic and cytokine assays

WT and patient derived DOCK8-expressing FV-infected NK-92 cells were independently mixed at effector to target ratios of 1:1 and 5:1 with K562 erythroleukemia cells for 4 hours as detailed previously.(20) For lytic assays, K562 target cells in the absence of NK-92 cells served as a background control. For degranulation (CD107a expression following 1:1 incubation for 0-2 hours with K562 cells) assays, NK-92 cells in isolation served as a background control. Target cell lysis and NK-92 cell degranulation were measured by flow cytometry (FCM) (LSRFortessa, BD Biosciences, Franklin Lakes, NJ) using live/dead fixable cell dead reagent (Invitrogen, Waltham, MA, USA) and anti-CD107a/LAMP1 allophycocyanin (APC)-conjugated antibody (Biolegend, San Diego, CA, USA), respectively, and analyzed with FlowJo 10.2 software (Ashland, OR, USA).(20) FCM intracellular detection of tumor necrosis factor (TNF) and interferon-gamma (IFNγ) of WT and patient derived DOCK8-expressing FV-infected NK-92 cells incubated 1:1 with K562 cells for 4 hours was carried out as previously detailed,(17) using fluorochrome-conjugated anti-cytokine antibodies (anti-IFNγ-APC, PharMingen, San Diego, CA, USA; anti-TNF-phycoerythrin (PE), Biolegend), anti-granzyme B-APC, and isotype controls (eBioscience, San Diego, CA, USA) in the presence of brefeldin A (golgi retention) and saponin (permeabilization).(22) K562 cells were labeled with eFluor 670 dye (Invitrogen, Waltham, MA).

### Splice variant reporter gene constructs

The modified EGFP reporter plasmids were constructed using traditional cloning methodologies as follows: protocol one (developed novel assay), the EGFP cDNA was spliced into the equivalent of 2 in-frame “exons” – EGFP 1/2 (1-336 bp) and EGFP 2/2 (337-720 bp); a centrally truncated DOCK8 intron 1 (416 bp, containing the 5’ and 3’ regions of the intron) – either WT 3’ splice acceptor site variant or patient derived mutation was inserted between the EGFP “exons”, as shown in Fig 2, A. For protocol two (standard exon trapping plasmid assay), the first 2 DOCK8 exons (53 bp and 103 bp in length, respectively) plus the EGFP cDNA serve as multiple exons; shortened introns from DOCK8 (either WT or mutant 3’ splice acceptor site of DOCK8 intron 1, 359 bp, and DOCK8 intron 2, 783 bp in length) were inserted in frame between the 3 exons, as shown in Fig 2, D. The plasmids were electroporated into NK-92 cells using a 4D-Nucleofector (Lonza, Basel, Switzerland) according to the manufacturer’s instructions. GFP expression was detected by FCM.

**FIG 1.**
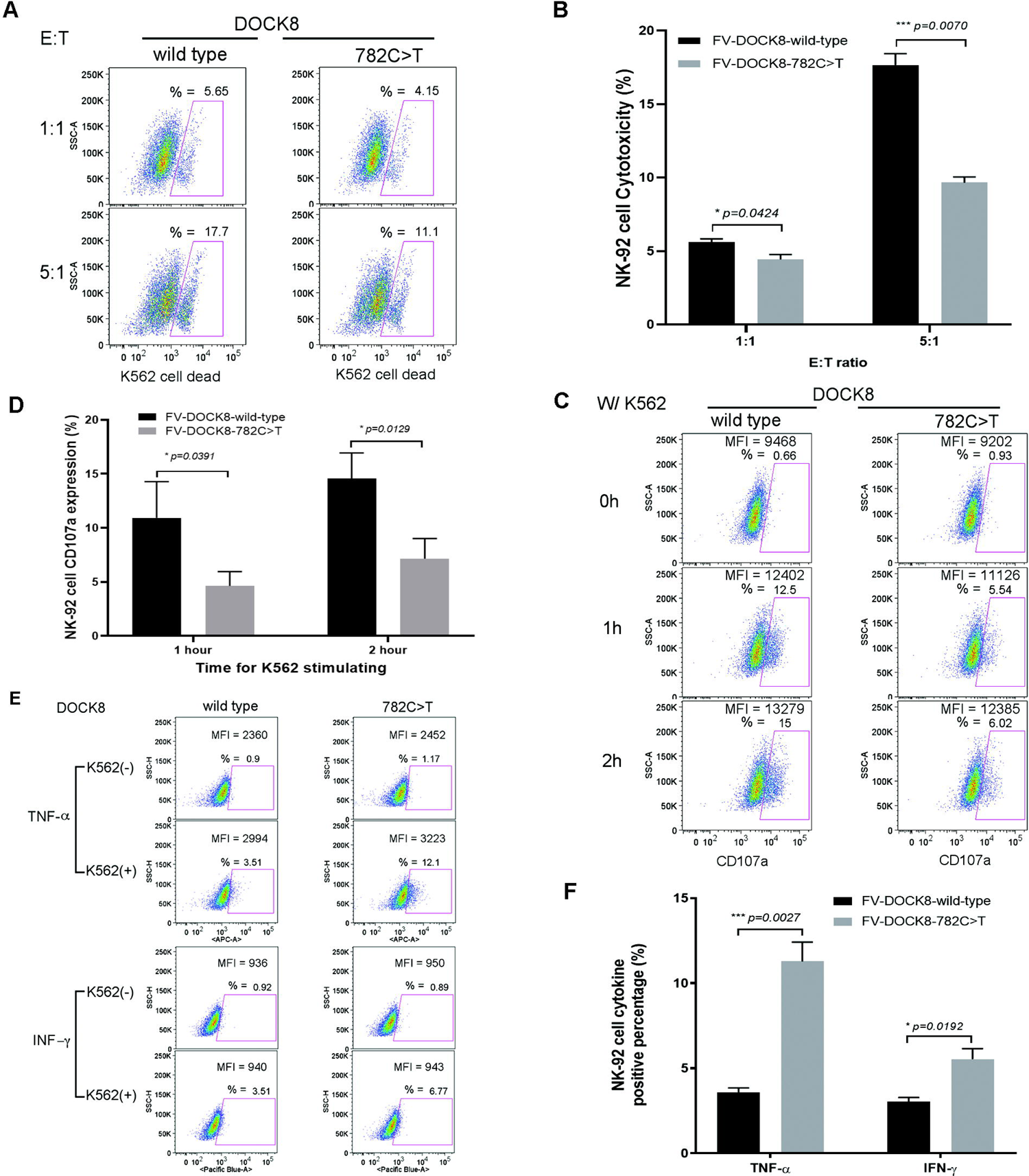
Over-expression of CSS patient 1 DOCK8 mutation diminishes NK cell function. Representative flow cytometry plots (**A**, **C**, **E**) and summary bar graphs (**B**, **D**, **F**) with means ± s.d. (n=4) and p values, showing decreased cytotoxicity (**A**, **B)**, decreased degranulation (CD107a expression) (**C**, **D)**, and increased intracellular TNF and IFNγ expression (**E**, **F)** when NK-92 cells were transfected with FV expressing DOCK8 c.782C>T compared to WT DOCK8.

**FIG 2.**
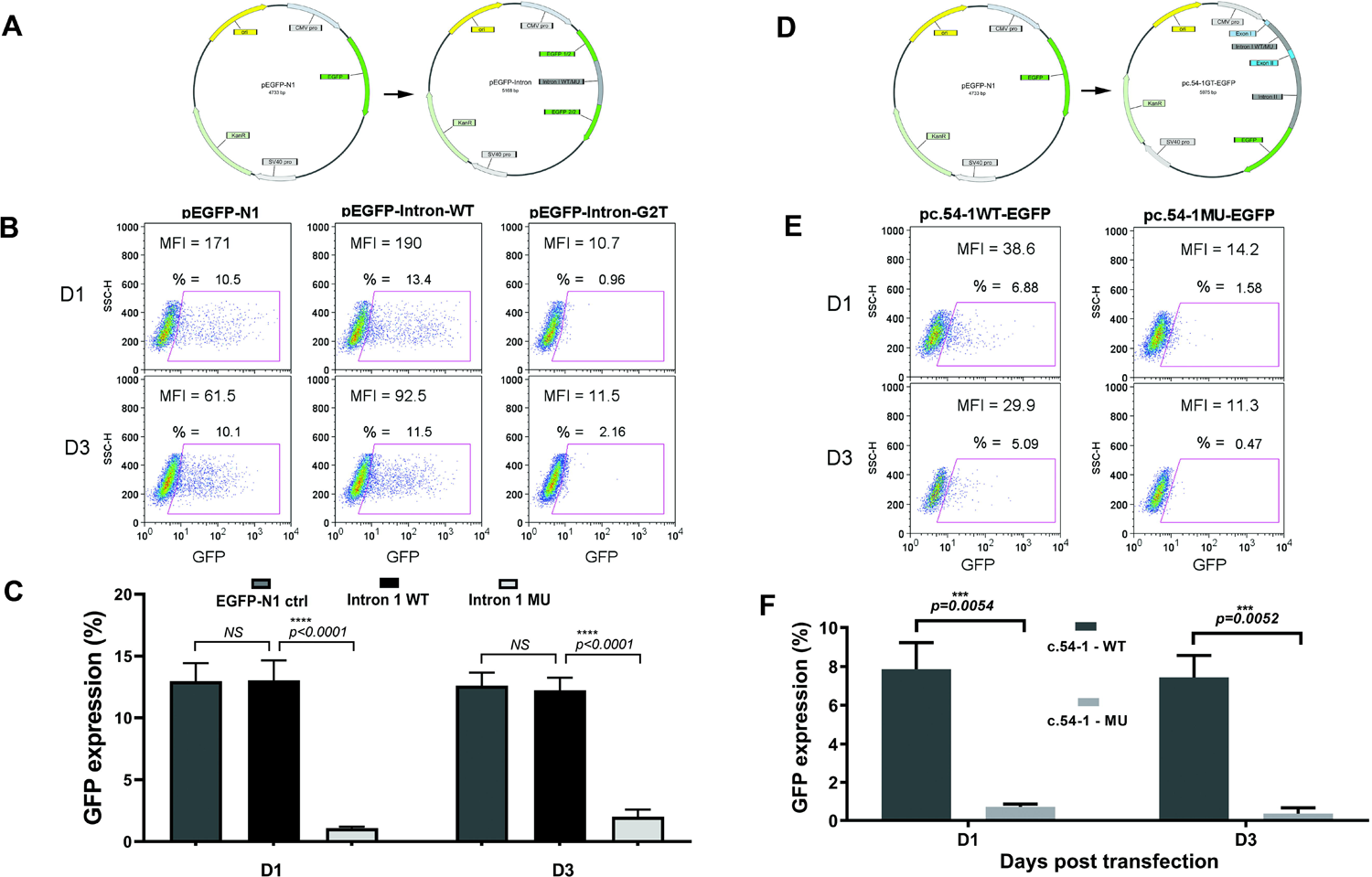
CSS patient 3 derived DOCK8 splice acceptor site nucleotide mutation in DOCK8 intron 1 disrupts RNA splicing in NK-92 cells. A. Representations of EGFP-expressing plasmid constructs pEGFP-N1 (left) and pEGFP-Intron (right) with DOCK8 intron 1 sequence disrupting the EGFP cDNA. **B.** Representative flow cytometry plots of NK-92 cells expressing EGFP 1 day (top) and 3 days (bottom) following transfection with pEGFP-N1 (left, comparator), pEGFP-Intron-WT (middle, WT splice acceptor site control), and pEGFP-Intron-G2T with the patient derived DOCK8 splice acceptor site mutation (right, c.54-1G>A). **C.** Bar graph summary with EFGP expression means ± s.d. (n=4) and p values at day 1 and day 3. **D/E/F** A traditional (exon trapping, see Methods) complementary EGFP reporter plasmid approach for assessing RNA splicing is shown to the right.

### Small interfering RNA (siRNA) studies

For DOCK8 mRNA knockdown, the indicated amounts (Fig 3: 500 nM total per condition, with increasing amounts of DOCK8 siRNA relative to control siRNA) of 3-Dicer substrate DOCK8 siRNAs mixture (TriFECTa DOCK8 siRNA kit, purchased from Integrated DNA Technologies, IDT, San Diego, CA) were nucleofected (Lonza) into 2 x 10^6^ NK-92 cells.(23) Dose-dependent diminished DOCK8 mRNA levels were confirmed with ∼90% reduction of DOCK8 expression at maximal DOCK8 siRNA concentration compared with the negative control as detected by qPCR at 24 hours after transfection (data not shown). The transfected cells were used to test for cytolysis and degranulation 24 hours post-transfection as described above.

**FIG 3.**
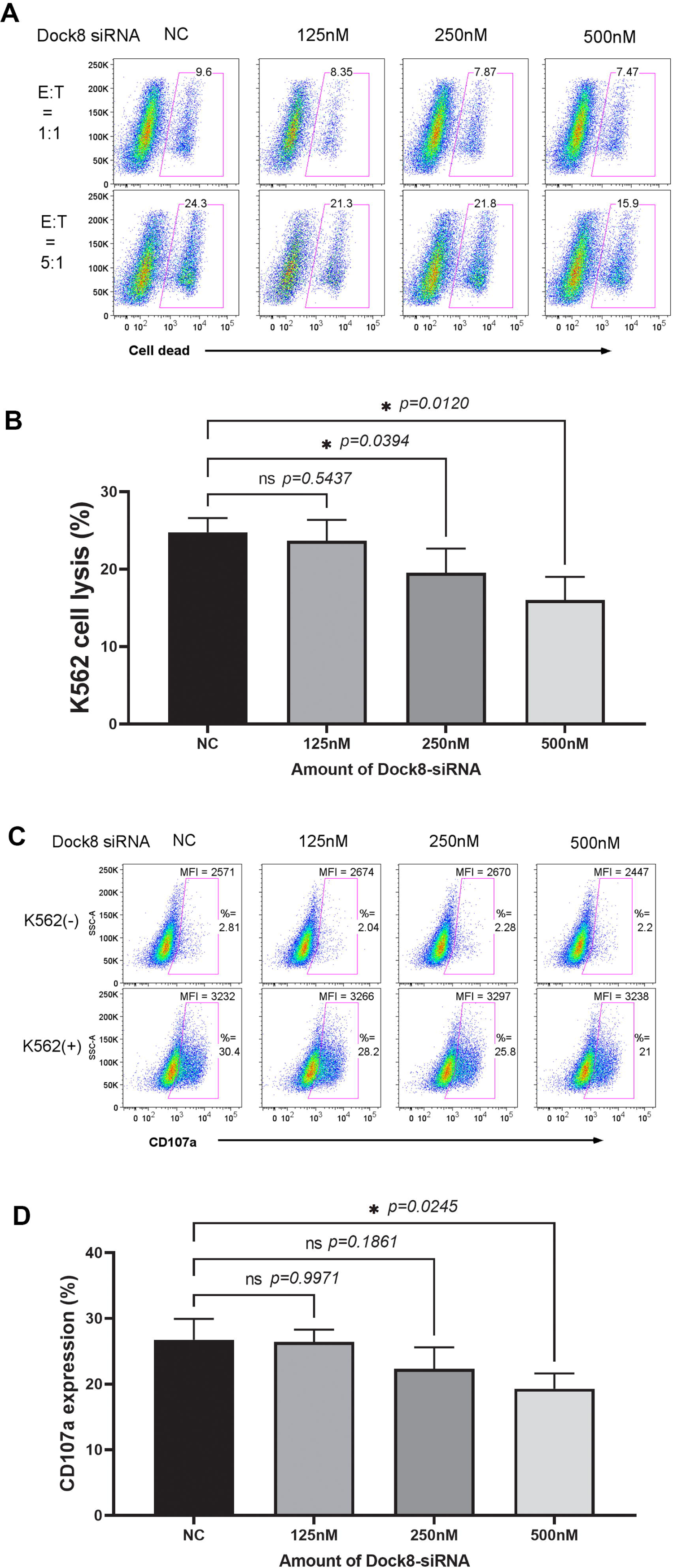
Stepwise knock-down of endogenous DOCK8 with siRNA yields decreased NK-92 cell cytotoxicity and degranulation. NK-92 cells were transfected with increasing amounts of DOCK8-specific siRNA (or scrambled sequence control, NC). The NK-92 cells were incubated with K562 target cells and representative flow cytometry plots depicting cytolysis (**A**) at 1:1 and 5:1 effector to target cell ratios and summary bar graphs (**B**) with mean cell lysis ± s.d. (n=3) and p values are shown. Similarly, DOCK8-siRNA transfected NK-92 cells were assessed for degranulation (CD107a) in the presence or absence of stimulatory K562 target cells. Representative flow cytometry plots (**C**) and summary bar graphs (**D**) with mean CD107a expression ± s.d. (n=4) and p values are shown.

### CRISPR/Cas9 genetic alterations

DOCK8 genomic editing was performed by using CRISPR(cr)/Cas9 technology.(24) Briefly, DOCK8 specific crRNA targeting the sequences (150 bp) in the vicinity of the patient 1 mutation (c.792T) was designed by using the IDT online service tool (CRISPR/Cas9 guide (g)RNA checker) and purchased from IDT. The most efficient, most specific, and with least off-target gRNA sequences were chosen (gel electrophoresis, data not shown). The crRNA and tracrRNA were first annealed to form the gRNA–ctRNA complex, followed by addition of Cas9 (Cas9-3NLS-v2, from IDT) to form the complete complex of Cas9–gRNA–ctRNA. The complex was nucleofected into NK-92 cells using a 4D-neucleofector (Lonza) according to the manufacturer’s instructions. An 88 bp enhancer oligonucleotide or 98 bp single stranded template oligonucleotide (ssODN, purchased from IDT) was also used during the transfection. Targeting of DOCK8 genomic DNA was confirmed by gel electrophoresis (data not shown). Transfected NK-92 cells were then studied for cytolysis and degranulation in the presence of K562 cells as described above.

### Mice studies

WT C57BL/6J mice were obtained from Jackson Laboratories (Bar Harbor, ME, USA). *Dock8*^-/-^ and *Il18tg* mice were generous gifts from Dr. Helen Su (National Institutes of Health, Bethesda, MD, USA)(25) and Dr. Tomoaki Hoshino (Kurume University),(26) respectively. All experimental mice were age matched. All experiments were conducted in accordance with national guidelines and approved by the Institutional Animal Care and Use Committee (IACUC) of the Children’s Hospital of Philadelphia. For LCMV infection, 2 x 10^6^ PFU of Armstrong strain LCMV was injected intraperitoneally into mice ages 6-8 weeks. Mice were weighed daily after infection. Cheek bleeding was used to obtain peripheral blood on day 0 and day 8 of infection for complete blood count analysis. Mice were euthanized on day 8. Spleens and livers were harvested for downstream analysis, and serum was obtained. For peripheral blood counts, 200 μL of peripheral blood was analyzed on a Sysmex XT-2000iV Hematology Analyzer (Kobe, Hyogo, Japan) to obtain white blood cell, red blood cell, and platelet counts. For detailed white blood cell analysis, red blood cells were lysed with ammonium-chloride-potassium (ACK) buffer, leukocytes stained with antibodies against B220, CD90, Ly-6G, NK1.1, TCRβ, CD4, CD8, and PD-1 (BD Pharmingen), cells assessed by flow cytometry, and data analyzed in FlowJo v10 (Treestar). Serum soluble CD25 (sCD25) and IFNγ were measured by ELISA using OptEIA kits (PharMingen) according to manufacturer’s protocol. Liver histology sections were fixed in formalin, embedded in paraffin, and stained with hematoxylin and eosin. A blinded pathologist (PAK) read the slides for lobular and portal inflammation, and endothelialitis according to published methods and scoring scales.(27)

### Statistics

Statistical analyses were performed with GraphPad Prism (GraphPad Software, La Jolla, CA, USA) software as described in figure legends.

## RESULTS

### Case presentation

A teen male (Table 1, patient 1), presented to the hospital with fever, headache, vomiting, non-bloody diarrhea, and abdominal pain for 4 days preceded by upper respiratory tract symptoms for 2 days. He developed acute respiratory distress syndrome (ARDS) requiring mechanical ventilation. Hepatosplenomegaly was noted on exam, and laboratory findings revealed bicytopenia, liver dysfunction, coagulopathy, hyperferritinemia, elevated sCD25, and greatly diminished NK cell lytic function (Table 2). The patient satisfied 7 out of the 8 HLH-2004 criteria(19) and met the HScore threshold(18) to diagnose HLH. An extensive infectious workup revealed *Bartonella henselae* infection by serology. Genetic sequence analysis showed a heterozygous novel mutation in the DOCK8 gene (c.782C>T, p.Ala261Val) and was negative for FHL genes. DOCK8 A261V is a conservative amino acid substitution, but *in silico* analysis predicts this variant is damaging to the protein structure/function. The mutation was predicted to be likely pathogenic by PolyPhen-2 (Table 1).

**Table 2.** HLH diagnostic features of patients with DOCK8 mutations.

| <u>Criterion/patient</u> | <u>1</u> | <u>2</u> | <u>3</u> |
| --- | --- | --- | --- |
| Underlying immunosuppression | No | Yes | Yes |
| Fever (°F) | 103.2 | 103.8 | 104.9 |
| Organomegaly (liver/spleen) | hepatosplenomegaly | none | splenomegaly |
| Cytopenia |  |  |  |
| Neutrophil count (<1E9/L) | 0.15 | 4.52 | 0.00 |
| Platelet count (<100E9/L) | 48 | 176 | 2 |
| Hemoglobin (<90 g/L) | 102 | 91 | 47 |
| Aspartate aminotransferase (≥30 U/L) | 267 | 520 | 134 |
| Triglyceride (≥265 mg/dL) | 329 | 253 | 1,044 |
| Fibrinogen (<1.5 g/L) | 3.6 | 2.4 | 5.1 |
| Ferritin (≥500 ng/mL) | 1,168 | 3,706 | 29,109 |
| NK cell activity (≤2.6 lytic units) | 0.6 | ↓CD107a | ND |
| Soluble CD25 (≥2,400 U/mL) | 5,187 | 2,075 | 12,081 |
| Hemophagocytosis | not in marrow | ND | present in marrow |
| HLH criteria met (≥5 of 8) | 7 | 2 | 7 |
| HScore (>169) | 209 (88-93%) | 195 (80-88%) | 292 (<99%) |
| Responsive to anakinra (rhIL-1Ra) | not given | yes | yes |

CD25 = interleukin-2 receptor alpha chain; HLH = hemophagocytic lymphohistiocytosis; HScore = reactive hemophagocytic syndrome diagnostic score; ND = not done; NK = natural killer; rhIL-1Ra = recombinant human interleukin-1 receptor antagonist

Two additional patients with HLH diagnosed by HLH-2004 and/or HScore criteria were identified with heterozygous DOCK8 mutations and no variants in FHL genes. Patient 2 (Table 1) was a teen boy with juvenile idiopathic arthritis (JIA, enthesitis related arthritis (ERA) subtype) with a rare DOCK8 missense mutation (c.4850A>G, p.Gln1617Arg) considered to be possibly pathogenic. Patient 3, a teen male, developed HLH (7/8 HLH-2004 criteria) (Table 2) in the setting of ongoing treatment for T-cell leukemia. He was noted to have a DOCK8 splice acceptor site mutation (c.54-1G>T) that was considered likely pathogenic (Table 1). All 3 HLH patients with heterozygous DOCK8 mutations survived their hospitalizations, with patient 1 responding to antibiotics and patients 2 and 3 responding to IL-1 blockade (anakinra) and glucocorticoids.

### Heterozygous HLH patient-derived DOCK8 missense mutations partially disrupt NK cell function

To explore the role of the heterozygous DOCK8 mutations from patient 1 and patient 2, a lentiviral expression construct was generated that co-expressed EGFP and WT (control) or mutant (c.782C or c.4850A) DOCK8 cDNA. Human NK-92 cells, which natively express both WT DOCK8 and the machinery for perforin-mediated cytolysis(17), were transduced with DOCK8 expressing WT or patient-derived FV as in the Methods. NK cell lytic activity of the DOCK8 c.782T (patient 1) expressing NK-92 cells was approximately half that compared to WT DOCK8 (c.782C) expressing cells. (Fig 1, A and B). Similarly, degranulation by DOCK8 c.782T expressing NK-92 cells was nearly half that of WT NK-92 cells (Fig 1, C and D). The decreased killing was not due to decreased granzyme B levels (Fig E1), but perhaps related to an appreciable decreased level of conjugate formation between NK-92 cells and the K562 target cells at later time points (30-60 minutes) post-mixing (Fig E2), as has been noted previously in DOCK8 deficiency.(28) Similar less robust, but statistically significant, reduction in NK cell lysis and degranulation was noted for the patient 2 derived DOCK8 c.4850A>G mutation (Fig E3, A-D). Thus, the FV introduced patient derived DOCK8 mutations function as partial-dominant negatives (in the presence of NK-92 WT DOCK8 germline genes) to inhibit NK cell lytic function.

We and others have previously shown the complete absence or partial disruption of genes in the perforin-mediated cytolytic pathway leads to delayed granule polarization to the immunologic synapse, diminished or absent killing, and prolonged engagement between the lytic lymphocyte (NK cell or CD8 T-cells) and its target cell.(17, 29, 30) The prolonged engagement is associated with increased pro-inflammatory cytokine production believed to contribute to the HIS.(17, 29, 30) NK-92 cells expressing WT (c.782C) or the patient 1 derived DOCK8 mutation (c.782T) were studied for intracellular cytokine expression following incubation with K562 target cells. NK-92 cells expressing the DOCK8 c.782T mutation produced approximately 3-fold more intracellular TNF and 2-fold more intracellular IFNγ compared to WT cells (Fig 1, E and F). This resembles the increased IFNγ production seen by a partial dominant-negative heterozygous RAB27A mutation identified in 2 unrelated teenagers with sHLH.(17)

### A HLH patient-derived splice acceptor site mutation disrupts DOCK8 mRNA splicing

The heterozygous mutation in DOCK8 (c.54-1G>T) from HLH patient 3 (Table 1) was in a predicted spice acceptor site at the 3’ end of intron 1. To explore the effect of the mutation functionally, we developed a novel plasmid-based assay. Using standard cloning techniques, we modified a standard EGFP expression plasmid (see Methods) in which the EGFP cDNA was interrupted in frame by inserting DOCK8 intron 1 possessing the WT (c.54-1 G) or the patient derived mutation (c.54-1 T) at the 3’ splice acceptor site, as if the EGFP cDNA was 2 exons separated by an intron. Thus, correct splicing out of intron 1 from DOCK8 would be required to generate EGFP RNA and subsequent protein to be detected by FCM.

Experimentally, EGFP expression plasmids were introduced into NK-92 cells by transfection, and the cells were placed back into culture. EGFP protein expression was screened by FCM at 1- and 3-days post-transfection. NK-92 cells transfected with the EGFP expression plasmid containing the EGFP cDNA disrupted by the DOCK8 intron 1 WT sequence (Fig 2, A) expressed similar levels of EGFP as the intact parent EGFP expression plasmid (Fig 2, B), but NK-92 cells transfected with the EGFP expression plasmid containing the EGFP cDNA disrupted by the DOCK8 intron 1 patient 3-derived mutation at the 3’ splice acceptor site (Fig 2, A) had minimal EGFP expression (Fig 2, B). Averaged over 4 separate experiments, NK-92 cells transfected with the WT DOCK8 intron 1 expressed roughly equivalent levels as those cells expressing the parent EGFP expression plasmid (Fig 2, C), but NK-92 cells transfected with the EGFP plasmid containing the DOCK8 intron 1 patient 3-derived mutation at the 3’ splice acceptor site had approximately 80-90% reduction in GFP expression (Fig 2, C). This argues that the patient-derived mutation at the 3’ end of DOCK8 intron 1 is required for optimal mRNA splicing and subsequent DOCK8 protein expression.

We confirmed these results by the more traditional and established plasmid-based exon trapping assay,(31) in which DOCK8 exon 1 and 2, separated by WT or patient mutation intron 1, were subcloned in frame and upstream of EGFP cDNA to provide more representative mRNA splicing context (Fig 2, D). Similar to the results of our own EGFP plasmid-based assay, NK-92 cells expressing the DOCK8 intron 1 mutant splice acceptor nucleotide had drastically diminished EGFP expression compared to those expressing the WT nucleotide (Fig 2, E and F). Thus, it is possible that HLH patient-derived heterozygous mutations may contribute to altered NK cell function by dominant-negative (Fig 1) or potentially hypomorphic effects (Fig 2).

### Decreased DOCK8 expression diminishes NK cell cytolytic function

To explore the effect of diminished DOCK8 expression on NK cell function, two complimentary approaches were undertaken. First, increasing amounts of DOCK8-specific siRNA were introduced into NK-92 cells by transfection. NK-92 cells were then assayed for lytic activity versus K562 target cells, and degranulation in the presence of stimulating K562 cells. Relative to scrambled siRNA control, increasing quantities of DOCK8-targeted siRNA reduced both killing and degranulation in a dose-dependent fashion as detected by FCM (Fig 3, A and Fig 3, C, respectively). Averaged over 4 independent experiments, knockdown of DOCK8 message statistically significantly reduced NK cell lysis by approximately 40% (Fig 3, B) and reduced degranulation (CD107a expression) by approximately 30% (Fig 3, D) at the highest concentration of DOCK8 siRNA (80-90% reduction of DOCK8 siRNA by RT-PCR, data not shown).

As an alternative approach, DOCK8 message was disrupted by targeting genomic DOCK8 DNA, using DOCK8-specific guide RNA, within NK-92 cells by CRISPR/Cas9 technology. Disruption of DOCK8 on both alleles resulted in approximately 50% reduction in NK cell lytic activity (Fig 4, A) and degranulation (Fig 4, C) as detected by FCM in a statistically significant manner averaged over 4 independent experiments (Fig 4, B and D). This argues that DOCK8 is required for optimal NK cell lytic activity but is not absolutely required for killing/degranulation. Re-introduction of WT DOCK8 DNA (c.782C) by CRISPR/Cas9 restored both K562 lysis and degranulation, whereas introduction of the patient 1 DOCK8 mutation (c.782T) was no better at NK cell lysis/degranulation than disrupted DOCK8 (Fig 4). This provides further evidence the patient 1 DOCK8 missense mutation disrupts NK cell lytic function.

**FIG 4.**
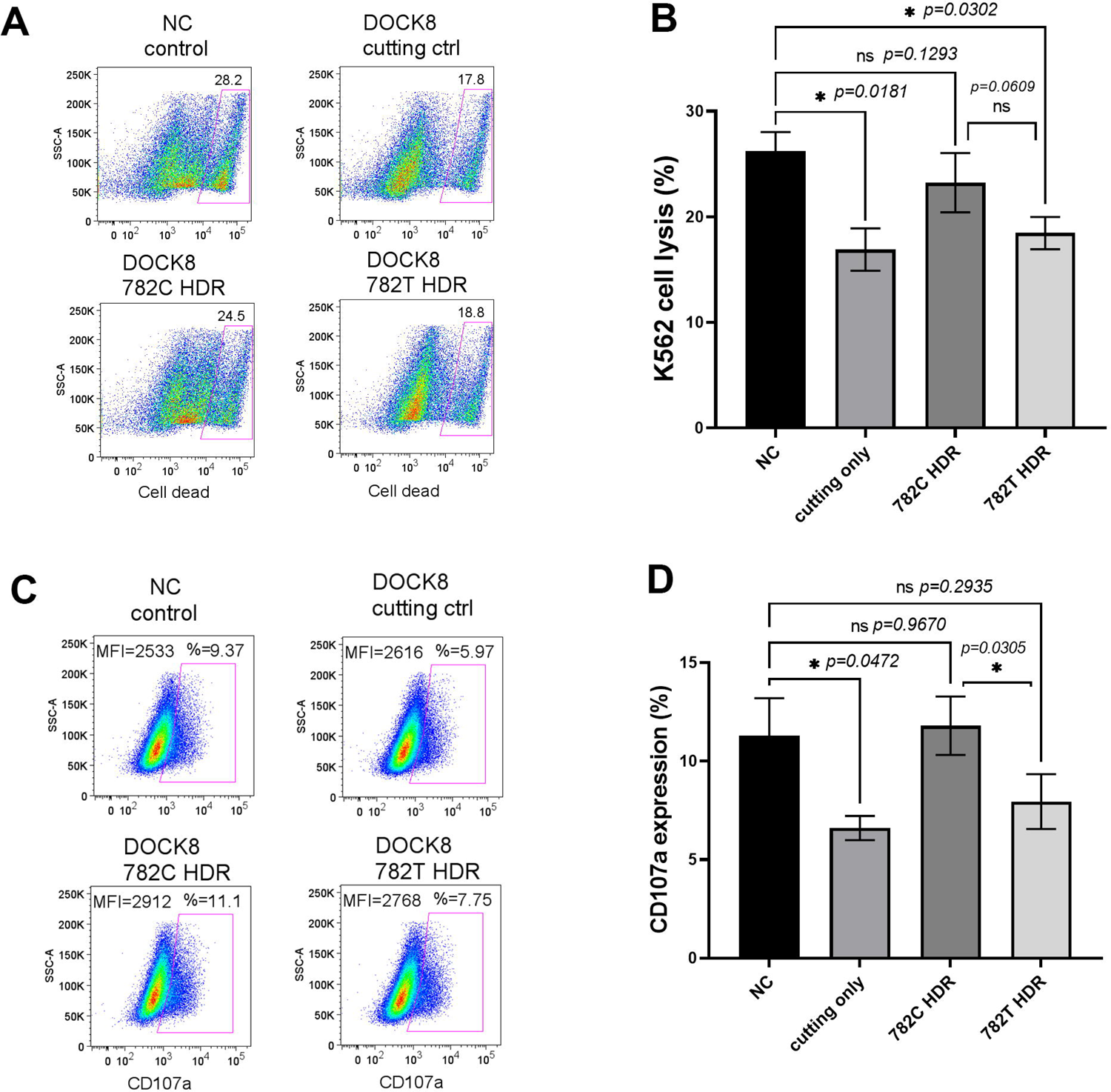
Knock-out and reintroduction of DOCK8 in NK cells with CRISPR/Cas9 demonstrates role of DOCK8 in NK cell cytolysis. Complete disruption (cutting ctrl) of DOCK8 in NK-92 cells by CRISPR/Cas9 diminishes NK-92 cell cytotoxicity (K562 cells) and degranulation (CD107a expressions). Reintroduction by homology-directed repair (HDR) of WT DOCK8 (782C), but not patient 1 mutation (782T), partially restores NK cell function. Representative flow cytometry plots of cell lysis (**A**) and degranulation (**C**) and summary bar graphs of mean cell lysis (**B**) and mean CD107a expression (**D**) ± s.d. (n=4) with p values are shown.

### LCMV infection of DOCK8 deficient mice leads to hyper-inflammation resembling HLH

To explore the role of DOCK8 in NK cell function *in vivo*, WT and *Dock8^-/-^* mice (32) were infected with LCMV as originally modelled in *Prf1^-/-^* mice.(33) After 4 days of infection, the B6 WT animals began to gain back body weight, whereas the *Dock8^-/-^* animals continued to lose weight until day 8 euthanization as required by protocol (Fig 5, A). As typically seen in humans and mice with HLH,(34) splenomegaly was noted in the LCMV infected *Dock8^-/-^*mice (Fig 5, B). Liver pathology was also noted, as often seen in human HLH, with increased endothelial activation (Fig E4). In addition, thrombocytopenia was noted in *Dock8^-/-^* mice relative to WT animals following LCMV infection (Fig 5, D), but no other relative cytopenias were identified (Fig 5, C and D). Serum IFNγ levels were slightly elevated in *Dock8^-/-^* mice compared to WT mice (Fig 5, E), similar but not as severe as in LCMV infected *Prf1^-/-^* animals.(33) Another common HLH feature (4) and marker of lymphocyte activation, elevation of serum sCD25, was not seen in *Dock8^-/-^* mice compared to WT controls (Fig 5, E). Lastly, and similar to *Prf1^-/-^* animals,(33) the *Dock8^-/-^*mice did not clear the LCMV infection as well in comparison to WT mice (Fig 5, F). Thus, consistent with *in vitro* disruption of DOCK8, in which DOCK8 disruption is not as severe as would be seen in PRF1 disruption (Fig 3 and Fig 4),(35, 36) LCMV infection of *Dock8^-/-^* mice leads to a hyper-inflammatory HLH-like state that is not as severe as in *Prf1^-/-^* animals.(33)

**FIG 5.**
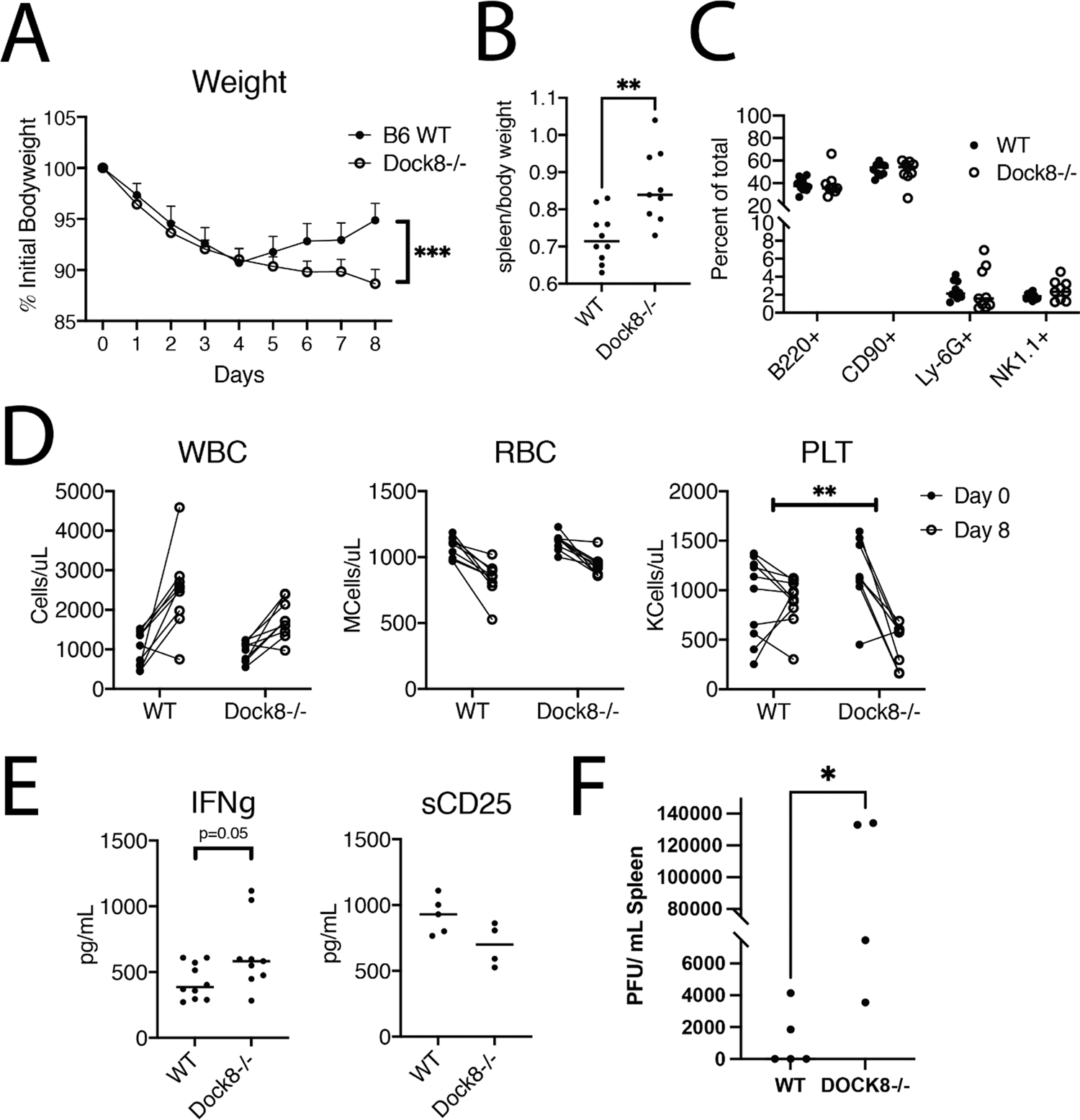
*Dock8*-/- mice infected with LCMV experience increased hyperinflammation relative to wild-type controls. Wild-type (WT) and DOCK8 deficient mice (*Dock8^-/-^*) were infected with 2 x 10^6^ PFU of LCMV intraperitoneally. **(A)** Daily weights expressed as % of initial body weight, error bars represent s.d. **(B)** Splenic weight normalized to total body weight, analyzed by t-test, ** – p <0.01. **(C)** Splenocytes were stained in multicolor flow cytometry for the listed markers, and percent of total splenocytes for each population is presented. **(D)** White blood cell count (WBC), red blood cell count (RBC), and platelet count (PLT) were analyzed from cheek bled peripheral blood pre-infection (day 0) and day 8 post infection. Counts were obtained using a Sysmex XT-2000iV Hematology Analyzer. Analyzed for genotype x disease interaction by two-way ANOVA, ** - p <0.01. b Serum IFNγ and sCD25 were measured by ELISA on day 8 post infection, analyzed by t-test. **(F)** Viral burden, analyzed by Vero cell plaque assay from splenic lysates, analyzed by t-test, * – p <0.05.

We recently have demonstrated synergy between cytotoxic impairment (perforin deficiency) and autoinflammation in the form of excess IL-18 (*Il18tg*) in driving spontaneous murine HLH.(37) To determine its interactions with excess IL-18, *Dock8*^-/-^ mice were crossed with *Il18tg* mice and studied for HLH features. *Dock8^-/-^*,*Il18tg* animals developed minimal evidence of spontaneous HLH, with trends towards increased spleen size and NK cytopenia (Fig 6, A). Using PD1 expression as a marker of CD8 T-cell hyperactivation,(38) *Dock8*^-/-^ mice showed CD4 and CD8 lymphopenia, whereas excess IL-18 rescued CD8 lymphopenia with an increase in activated CD8 T-cells (Fig 6, A). Similar, but more overwhelming, increases in hyperactivated CD8 T-cells were observed in human HLH/MAS(39, 40) and in *Prf1^-/-^;Il18tg* mice.(38) When infected with LCMV, *Dock8^-/-^;Il18tg* mice displayed slightly more weight loss, anemia, neutropenia, and thrombocytopenia (data not shown) than *Dock8*^-/-^ or *Il18tg* alone (Fig 6, B). Nearly all CD8 T-cells were hyperactivated (PD1+), but they were less abundant, likely reflecting Dock8-deficiency’s effect on CD8 T-cell survival (Fig 6, B).(41) The combination of the cytolytic impairment (*Dock8^-/-^*) and autoinflammation (*Il18tg*) in the setting of an infectious trigger (LCMV) supports the development of CSS using a threshold model of disease.(16)

**FIG 6.**
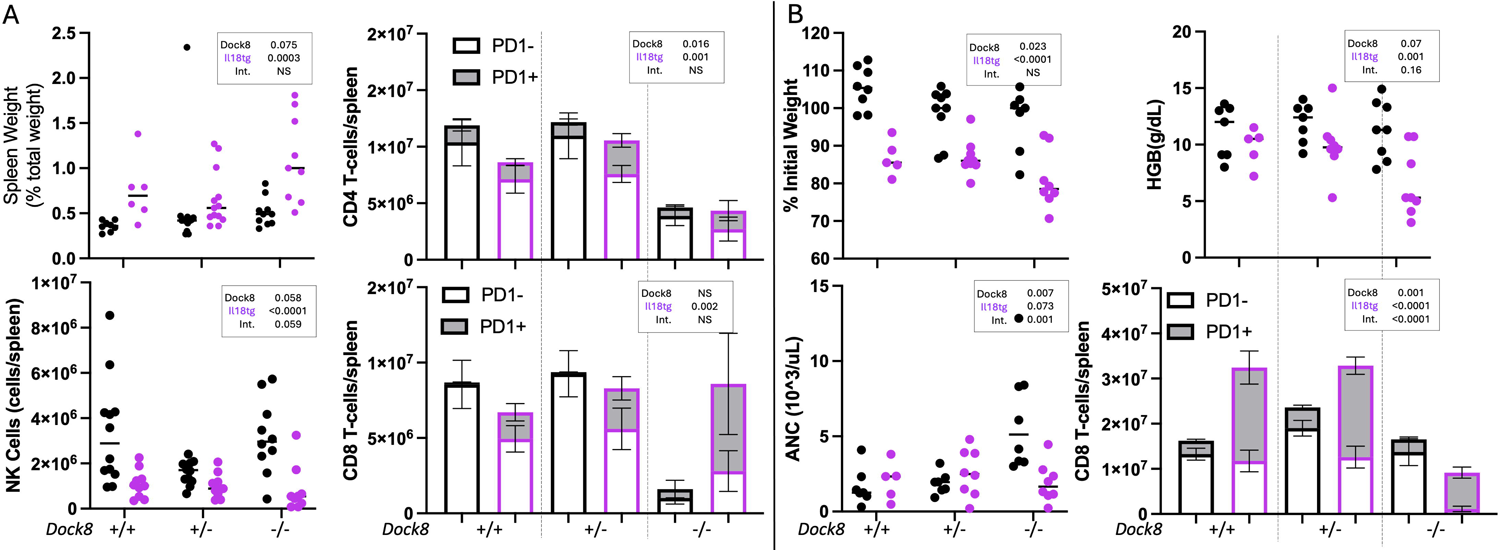
Despite driving T-cell lymphopenia, *Dock8* deficiency interacts with excess IL-18 to amplify CD8 T-cell activation and LCMV-induced HLH. Littermate mice of the indicated genotypes were assessed at 8-10 weeks of age for (**A**) spontaneous splenomegaly, splenic NK cell abundance, and the number and activation state of splenic CD4 and CD8 T-cells. Separately, mice of the same genotypes were infected with LCMV and assessed at Day 8-9 post infection for (**B**) weight loss, neutrophilia, anemia, and number and activation state of splenic CD8 T-cells. Data were analyzed by two-way ANOVA for significant differences based on *Dock8* genotype, presence of *Il18* transgene, and interaction. For bar graphs, ANOVA results indicate comparisons of the % of CD4 or CD8 T-cells that were PD1+. NS=not significant, indicating p>0.2.

## DISCUSSION

FHL typically presents in infancy resulting from homozygous defects in genes critical for perforin-mediated cytolysis.(42) Many (30-40%) of older children and adults with CSS, like HLH and MAS possess heterozygous defects in these well-established FHL genes(42) with partial or complete dominant-negative effects(43–45) leading to sHLH.(45, 46) However, there are CSS patients with defects in NK cell lytic function (e.g. patients 1 and 2, Table 1) without identified defects in known perforin pathway genes. We identified heterozygous DOCK8 mutations in 3 young patients with sHLH without perforin pathway gene mutations. Two of them (patients 1 and 2) displayed defective NK cell function (Table 2) and had novel or rare DOCK8 missense mutations (Table 1) resulting in partial defects in NK/cytotoxic T-cell killing (Fig 1 and Fig E3) likely contributing to the CSS in a partial-dominant negative fashion using a threshold model of disease.(6)

In such an additive threshold model, a genetic contribution to partial NK cell cytolytic function results in increased pro-inflammatory cytokine levels, which when added to the inflammation from an underlying condition, e.g. leukemia or systemic lupus erythematosus, and/or certain infectious triggers (e.g. EBV) may reach a threshold at which the host immune response can no longer tolerate the “cytokine storm” and contributes to the multiorgan system disruption.(6, 47) This model helps to explain how relatively subtle genetic defects in lymphocyte lytic function may normally be tolerated but contribute to CSS under certain conditions.(15) Homozygous genetic defects in perforin pathway genes require very little stimulus to overcome an immune balance threshold, but heterozygous defects that partially alter lymphocyte cytolysis may contribute to overcoming the CSS threshold in various heightened inflammatory settings.

Patient 1, with a heterozygous, likely pathogenic, and novel missense mutation in DOCK8 developed sHLH in the setting of an infectious trigger, *Bartonella henselae* (Table 1). The intracellular pathogen, Bartonella has been reported in association with sHLH in 2 cases with underlying renal transplantation.(48, 49) By comparison, patient 2, with a heterozygous, possibly pathogenic, rare missense DOCK8 mutation, developed CSS in the setting of a JIA/ERA disease flare (Table 1). Of note, t we have formerly reported 2 children who developed sHLH with partial dominant-negative mutations in RAB27A and UNC13D, respectively, who both went on to develop JIA/ERA.(17, 50) Thus, the threshold model of disease may help explain how partial defects in lymphocyte cytolysis from heterozygous missense mutations in DOCK8 (Fig 1 and Fig E3, respectively) contribute to sHLH development in the context of an intracellular pathogen (Table 1, patient 1) or an autoimmune disease flare (Table 1, patient 2). Recently, we reported 4 out of 39 children with the post-COVID-19 HIS, multisystem inflammatory syndrome in children (MIS-C),(51) who possessed unique DOCK8 missense mutations, all with partial dominant-negative effects on NK cell lytic function.(20) We are currently exploring the function of 3 unique DOCK8 missense mutations from 20 severe COVID-19 adults who underwent genetic sequencing as part of a clinical trial.(52)

In contrast to patients 1 and 2, the rare, likely pathogenic DOCK8 mutation from patient 3 (Table 1) yielded a splice acceptor site defect that disrupted RNA splicing (Fig 2). A heterozygous hypomorphic defect may or may not have contributed to CSS development in patient 3, who suffered from T-cell leukemia (a known HLH trigger)(53) and a potential Candida infectious trigger in the setting of iatrogenic immunodeficiency.(54) Nevertheless, heterozygous hypomorphic defects in FHL genes may contribute to sHLH development.(55) As DOCK8 does not appear to be absolutely required for NK cell lytic function (Fig 4), the contribution of a heterozygous hypomorphic mutation in DOCK8 to CSS may be difficult to establish. Nonetheless, peak DOCK8 levels appear to be required for optimal NK cell cytolytic activity (Fig 3). Moreover, a prior report details a heterozygous splice donor site mutation in DOCK8 contributing to impaired lymphocyte cytotoxicity.(56) Thus, heterozygous hypomorphic DOCK8 genetic defects may contribute to CSS in a threshold model of disease.(6)

By comparison, homozygous disruption of DOCK8, results in more severe defective NK cell lytic function in humans(11, 12) and homozygous DOCK8 deficiency was reported to be associated with HLH in a child.(9) Herein, we studied a murine model of FHL by infecting *Dock8^-/-^* mice with LCMV (Fig 5) and noted a HIS resembling, but not as severe as, LCMV infected *Prf1^-/-^* animals.(33) While LCMV infected *Dock8^-/-^* mice, in comparison to WT animals, did not clear infection well, and had features of sHLH (splenomegaly, thrombocytopenia, liver inflammation) (Fig 5 and Fig E4), they only had minimally elevated levels of IFNγ (Fig 5, E). Perhaps, this relates to the importance of DOCK8 in lymphocyte cytokine production,(11) such that there is a trade-off between a cytokine production defect(57, 58) and increased cytokine production resulting from prolonged interaction between defective lytic lymphocytes and their stimulating infected target cells.(29, 30) This may help explain why children with Hyper-IgE syndrome secondary to DOCK8 deficiency are not commonly reported to develop CSS.(59) Nevertheless, the addition of autoinflammation (*Il18tg*) on top of the cytolytic defect (*Dock8^-/-^*) contributes to a more severe HIS in the LCMV infection model of murine HLH (Fig 6).

The mechanism by which heterozygous DOCK8 missense mutations disrupt NK cell cytolytic activity is not entirely clear. Others have shown that disruption in DOCK8 has no effect on intracellular perforin levels,(57, 58) and we show that the patient 1 DOCK8 missense mutation has no effect on granzyme B levels (Fig E1) despite the partial dominant-negative effect on NK cell cytolytic activity (Fig 1). Others have reported that DOCK8 is not required for NK cell conjugation with its target cell(11), but our data suggest that a partial dominant-negative DOCK8 missense mutation may subtly diminish conjugate formation (Fig E2). The ability of DOCK8 to activate CDC42, together with Wiskott-Aldrich syndrome protein, is critical for coordinated F-actin reorganization and proper formation of the lytic immunologic synapse between NK cells and their target cells.(12) Thus, partial dominant-negative missense mutations in DOCK8 may delay cytolytic granule trafficking to the immunologic synapse as has been shown for a partial dominant-negative missense mutation in RAB27A identified in 2 unrelated children with sHLH. Exactly how a partial dominant-negative DOCK8 missense mutation would alter CDC42 (a newly described HLH related gene)(13) activation and subsequent actin-dependent cytolytic granule movement is unclear, as *in silico* modeling of the patient 1 and patient 2 DOCK8 missense mutations does not predict a direct disrupted interaction with CDC42 (data not shown) based on crystal structure data.(60) Nonetheless, both patient-derived DOCK8 missense mutations partially disrupt NK cell cytotoxicity (Fig 1 and Fig E3) and likely contributed to sHLH development.

## CONCLUSIONS

Herein, we describe 3 individuals with sHLH, all with heterozygous mutations in the primary immunodeficiency gene, DOCK8, which is well known to be required for optimal NK cell cytolytic function. We demonstrate the 2 DOCK8 missense mutations inhibit NK cell lysis and degranulation when introduced into NK-92 cells via FV. The other DOCK8 mutation, altering a spice acceptor site nucleotide, was shown to disrupt RNA splicing using a novel and an established reporter gene electroporated into NK-92 cells. Stepwise (siRNA) or complete disruption (CRISPR/Cas9) of DOCK8 in NK-92 cells substantially diminished NK cell lytic function but does not appear to be absolutely required. Lastly, LCMV infection of *Dock8^-/-^* mice resembles murine HLH but is not as severe as traditional FHL mouse models. Taken together, we propose that DOCK8 be considered a sHLH associated gene.

## Data Availability

All data produced in the present work are contained in the manuscript.

## ACKNOWLEDGMENTS

We thank the patients and their families for participation in these studies.

## Abbreviations used

CSS: cytokine storm syndrome

DOCK8: dedicator of cytokinesis 8

FV: foamy virus

HIS: hyper-inflammatory syndrome

HLH: hemophagocytic lymphohistiocytosis

IBD: inflammatory bowel syndrome

LCMV: lymphocytic choriomeningitis virus

MAS: macrophage activation syndrome

NK: natural killer

---

**FIG E1.**
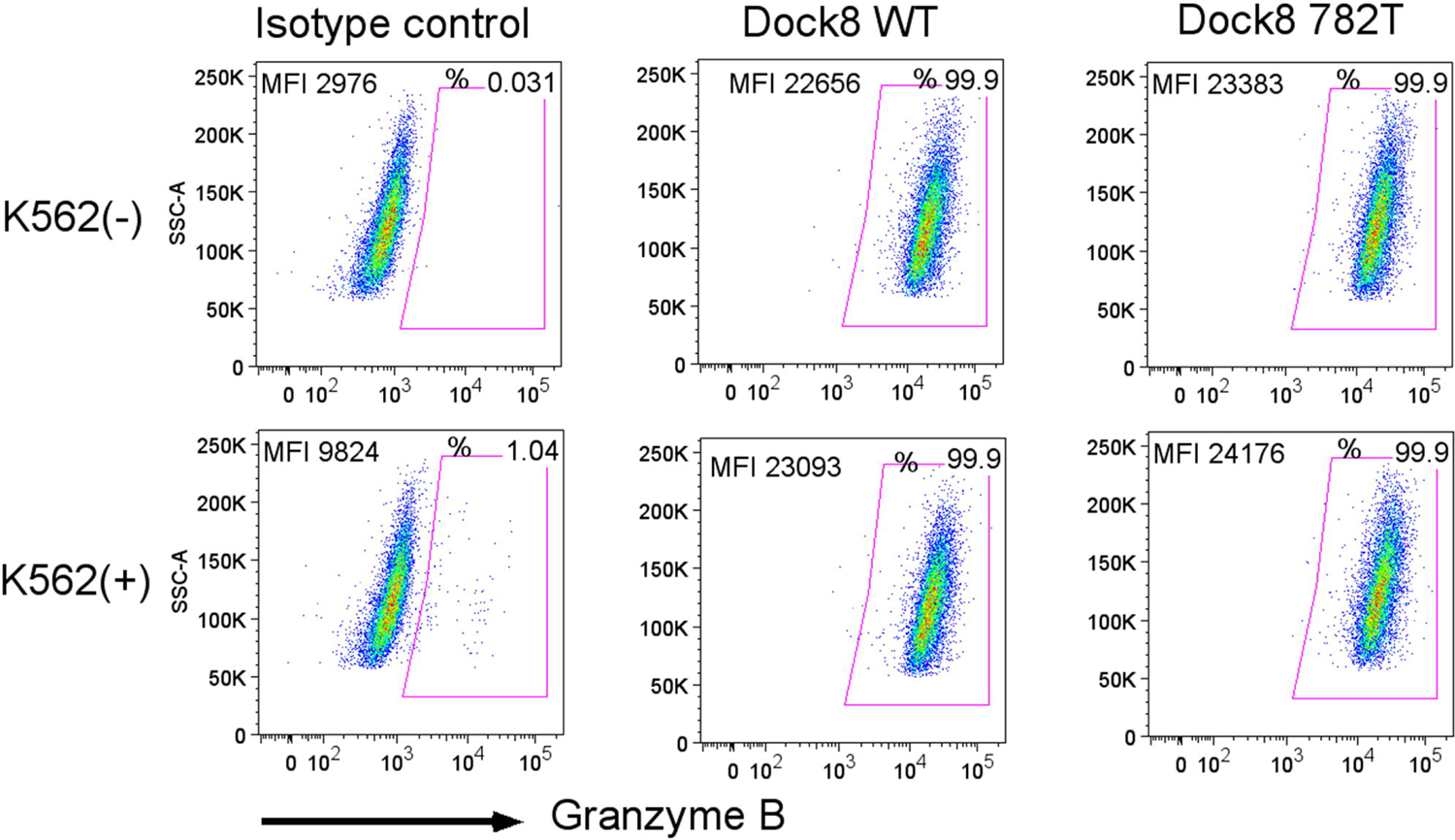
Patient 1 DOCK8 mutation has no effect on granzyme B expression. NK-92 cells were analyzed by FCM in the presence (+) or absence (-) of K562 cells for intracellular granzyme B expression (2^nd^ row from top) in comparison to APC-conjugated isotype control antibody (top row). WT (3^rd^ row from top) or DOCK8 mutant (c.782T) (bottom row) FV transfected NK-92 cells were assessed for granzyme B expression in a similar fashion. MFI and percentage positive cells are reported on each FCM plot (side scatter on y-axis, APC detection on x-axis), and data shown are representative of 4 independent experiments.

**FIG E2.**
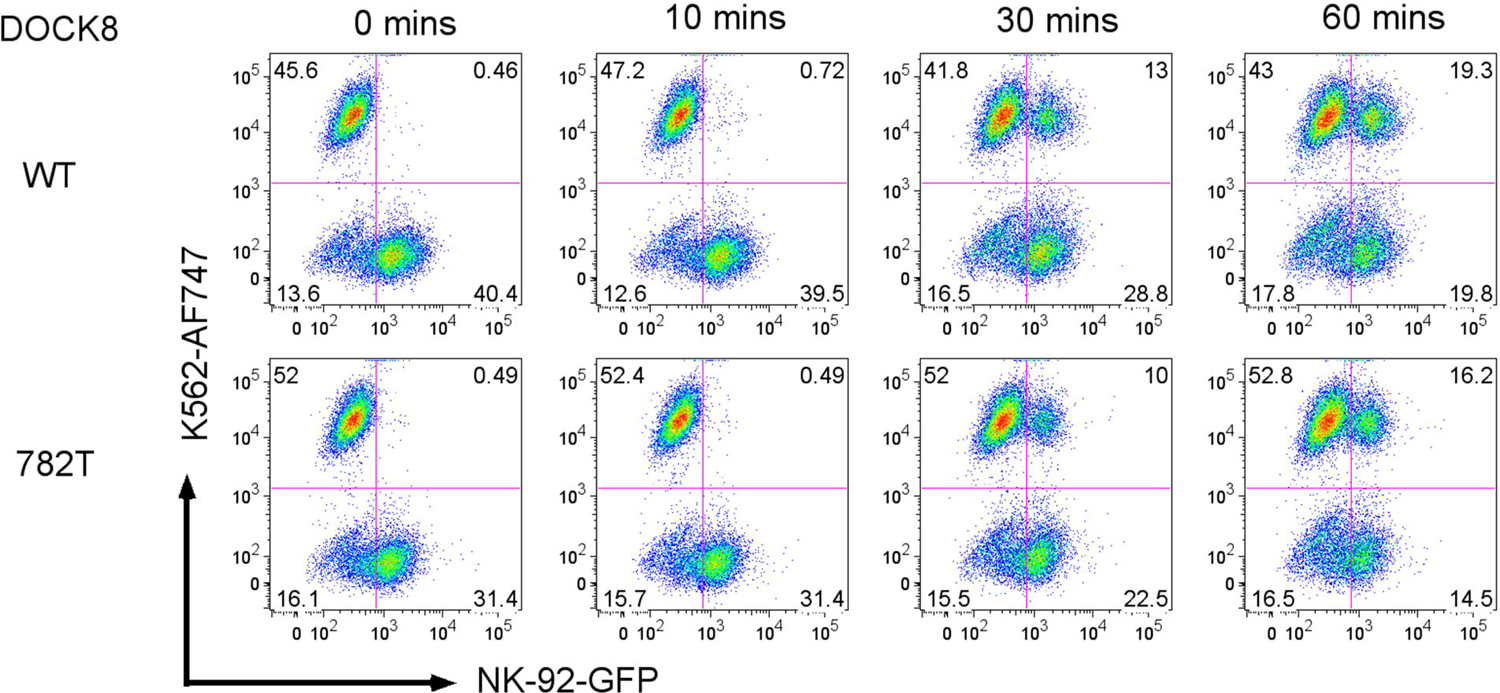
Patient 1 DOCK8 mutation modestly diminishes NK-92 cell and K562 cell conjugation. WT (c.782C) or patient 1 DOCK8 mutation (c.782T) FV transfected NK-92 cells (co-expressing EGFP, y-axis) were co-incubated 1:1 with K562 target cells (labeled with eFluor 670 and detected by APC emission wavelength, x-axis) for 0, 10, 30, or 60 minutes. FCM plots depicting NK-92 cells binding as a percentage of K562 cells (top rows), or K562 cells binding as a percentage of NK-92 cells are presented (middle rows). The upper right quadrants in the bottom rows show the percentages of NK-92 cell to K562 cell conjugates with quadrant percentages listed. Data shown are representative of 4 independent experiments.

**FIG E3.**
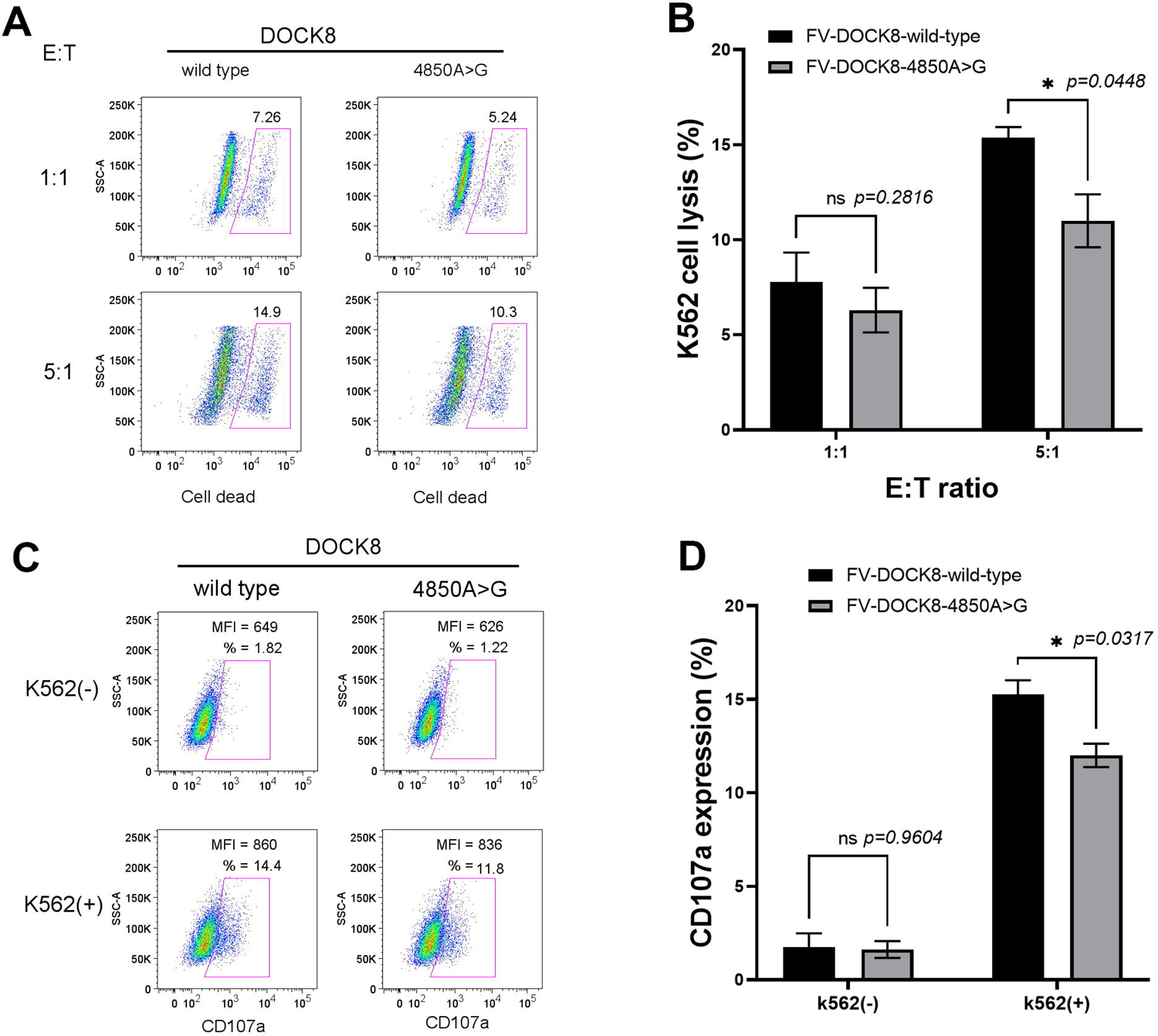
Over-expression of CSS patient 2 DOCK8 mutation diminishes NK cell function. Representative flow cytometry plots (**A**, **C**) and summary bar graphs (**B**, **D**) with means ± s.d. (n=4) and p values, showing decreased cytotoxicity (**A**, **B)** and decreased degranulation (CD107a expression) (**C**, **D)** when NK-92 cells were transfected with FV expressing DOCK8 c.4850A>G compared to WT DOCK8.

**FIG. E4.**
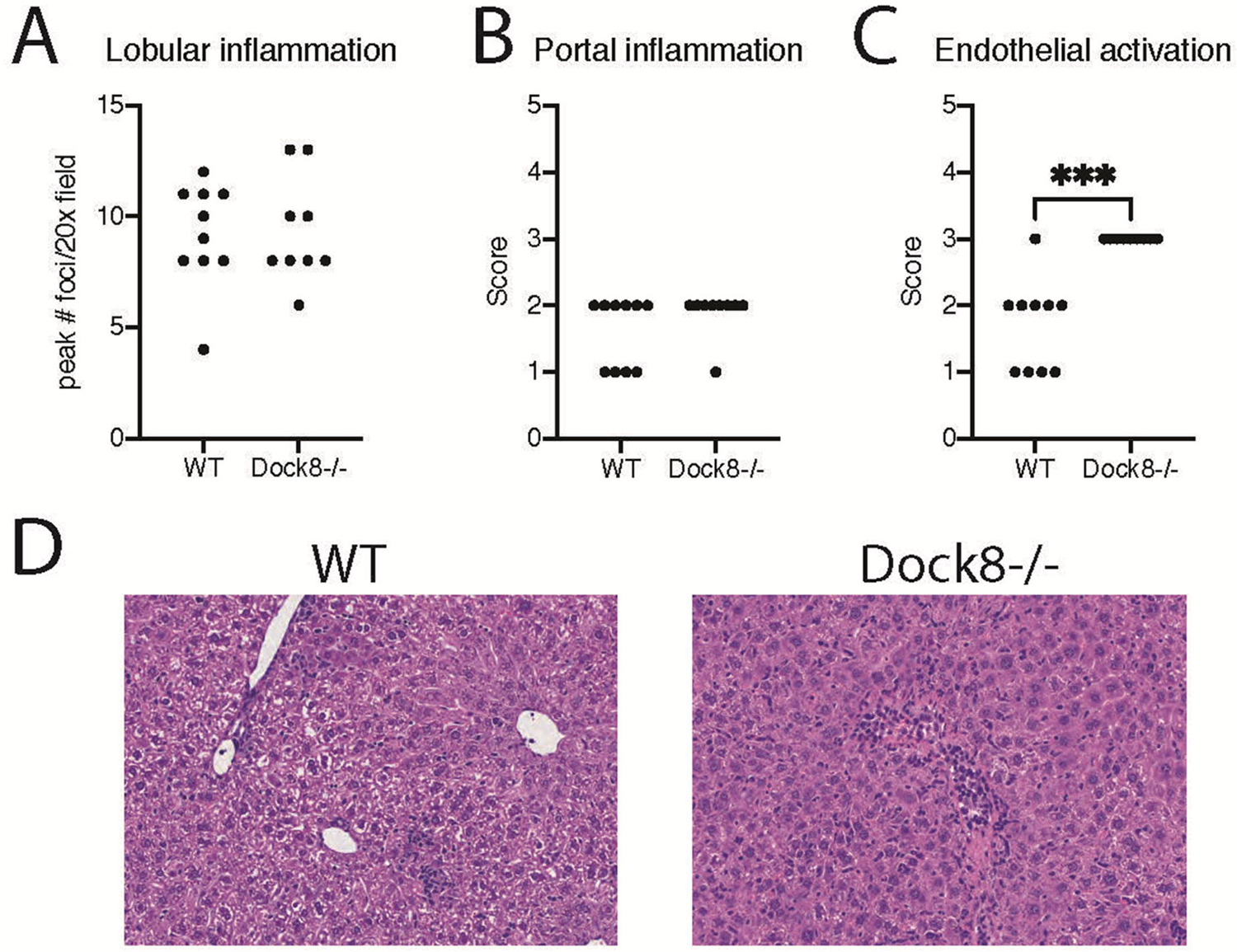
Pathology of *Dock8*^-/-^mice infected with LCMV demonstrate increased hepatic endothelial cell activation. Mice were infected with LCMV as in Fig 5. Livers were harvested on day 8 and were assessed by a blinded pathologist and scored for various features. **(A)** Lobular inflammatory score as measured by number of inflammatory foci per 100× field in the most involved area. **(B)** Portal inflammatory score as measured on a scale of 0-4 with 0 being absent and 4 representing severe inflammation. **(C)** Histologic endothelial activation as measured by morphologic changes in the vascular endothelial cells was 0-4 with 0 being normal and 4 representing severe changes of rounding of cells, plumpness of cytoplasm and enlargement of nuclei. **(D)** Representative histomicrographs of livers from WT and *Dock8^-/-^* mice showing complete occlusion of venules by inflammatory cells in *Dock8^-/-^* mice.

